# Cross-sectional study of the association between perceived organizational support and COVID-19 vaccine intention

**DOI:** 10.1101/2021.05.07.21256839

**Authors:** Yuichi Kobayashi, Tomohisa Nagata, Yoshihisa Fujino, Ayako Hino, Seiichiro Tateishi, Akira Ogami, Mayumi Tsuji, Shinya Matsuda, Koji Mori, CORoNaWork project

## Abstract

**Objectives:** This study examined the association between perceived organizational support (POS) and COVID-19 vaccine intention and the influence of the implementation of workplace infection prevention measures.

**Methods:** We analyzed 23,846 workers using data from an Internet survey of workers aged 20–65 years conducted in December 2020, during a period of widespread COVID-19 infection in Japan.

**Results:** A higher POS was associated with a higher intention to vaccinate. The relationship between POS and vaccine intention was attenuated when adjusted for infection prevention measures in the workplace.

**Conclusions:** In workplaces where POS is present, a sense of responsibility to the group and altruistic behavior may arise. This means employees act to acquire herd immunity to protect others, which may result in increased vaccine intention. The association between POS and vaccination intention was attenuated by adjusting for workplace infection prevention measures, which suggested that infection prevention measures may be a confounding factor or that POS created a health climate that promoted infection prevention measures. The results suggest that working to improve employee well-being and implementing appropriate workplace infection prevention measures during infectious disease outbreaks may promote vaccination behavior and contribute to the acquisition of herd immunity in the community.

## Introduction

On March 11, 2020, the World Health Organization declared the spread of coronavirus disease 2019 (COVID-19)^1)^ was a pandemic caused by the severe acute respiratory syndrome coronavirus 2 (SARS-CoV-2). As of April 1, 2021, 128,822,735 people worldwide had been infected with COVID-19 and 2,815,166 people had died^2)^. Although the number of infected people in Japan was lower than that in Europe and the United States, the Japanese government declared a state of emergency three times (as of April 25, 2021)^3)^. It is important to implement preventive actions against emerging infectious diseases, and actions such as keeping a physical distance, wearing masks, and hand disinfection were recommended to help combat COVID-19^4)^. In addition to these infection prevention measures, acquisition of herd immunity is important to end the pandemic^5)^. However, given the few infected people at that time, a survey conducted in December 2020 reported the antibody prevalence among Japanese people was 1.35% in Tokyo, which was the city with the highest prevalence^6)^.

The newly-developed COVID-19 vaccine is being administered in many countries to achieve herd immunity. It is also important to achieve herd immunity through vaccination in Japan to protect people from COVID-19 and restore economic activities. In Japan, vaccination of approximately 40,000 healthcare workers began on February 17, 2021, and vaccination of older people (aged ≥65 years) began in April 2021^7)^. However, many people are hesitant to be vaccinated, and some even refuse vaccination^8)^. This vaccine hesitancy is considered a major barrier to the acquisition of herd immunity against SARS-CoV-2. Vaccines may cause adverse reactions, and there are ethical issues involved in making vaccination mandatory. Therefore, in most countries, vaccination for COVID-19 is voluntary and based on individual decision making^9)^. The vaccination rate in Japan is lower than that in other countries, including for the seasonal influenza vaccine^10)^, influenza A (H1N1) that occurred in 2009^11)^, and the human papilloma virus vaccine (which appeared to be associated with side effects although the causal relationship was unclear)^12)^. Addressing vaccine hesitancy in Japan is therefore important to achieve herd immunity to COVID-19.

Organizational factors such as leadership in health promotion and the health climate have a significant impact on employees’ health behaviors^13)^. With regard to vaccination, in workplaces with a good health climate that supports employees’ health promotion and when infection prevention measures are actively implemented, the number of people in that workplace who are vaccinated may increase. Previous literature on organizational factors in the workplace and vaccination behavior reported that the presence of organizational governance influenced vaccination rates in healthcare organizations^14)^.

An indicator related to such a health climate is perceived organizational support (POS). POS is based on social exchange theory and refers to an employee’s own perception of whether their organization is supportive of improving the well-being of its employees. A previous study found that employees with a high POS were more likely to participate in wellness programs compared with those with a low POS^15)^. Therefore, employees working in organizations with high POS may be more willing to be vaccinated to help prevent infection in the workplace. Furthermore, during the COVID-19 pandemic, employers are expected to be proactive in adopting infection prevention measures in collaboration with their employees. This suggests that proactive infection prevention measures that seek to protect the health of employees may increase POS. However, no previous studies have examined the relationship between POS in the workplace during a pandemic outbreak or the status of infection control efforts in the workplace and vaccine intentions.

We therefore formulated two hypotheses: 1) “Employees with a high POS have a high intention to be vaccinated with the COVID-19 vaccine” and 2) “The association between POS and vaccine intention is influenced by the implementation status of infection prevention measures in the workplace.” We examined these hypotheses using data from an Internet survey conducted in Japan when COVID-19 vaccination was in the planning stage.

## Materials and Methods

A research group from the University of Occupational and Environmental Health, Japan, conducted a prospective cohort study, known as the Collaborative Online Research on Novel-coronavirus and Work study (CORoNaWork study). The self-administered questionnaire survey was completed by a panel registered with the Internet survey company Cross Marketing Inc. (Tokyo, Japan). During the baseline survey (conducted December 22–25, 2020), Japan was in the third wave of the pandemic in which the numbers of COVID-19 infections and deaths were markedly higher than in the first and second waves; therefore, the country was on high alert.

We used baseline survey data from the CORoNaWork study to conduct the present cross-sectional study. The CORoNaWork study protocol, including the sampling plan and participant recruitment procedure, has previously been reported in detail^16)^. Participants were aged 20–65 years and working at the time of the baseline survey (N = 33,087), and stratified using cluster sampling by gender, age, region, and occupation. After excluding 6,051 participants who provided invalid responses, we included data for 27,036 participants from the database. The exclusion criteria for the present study were: individuals who had been infected with the COVID-19 or who had been a close contact of a person who was diagnosed with COVID-19; self-employed workers; workers in small/home offices; and agriculture, forestry, and fishery workers. We finally analyzed data for 23,846 workers.

The present study was approved by the Ethics Committee of the University of Occupational and Environmental Health, Japan (reference No. R2-079). Informed consent was obtained through the CORoNaWork study survey website at the time data were collected.

### Assessment of POS

POS was evaluated with the following question. “Your company supports employees in finding a balance between active, productive working and healthy living,” based on a previous study^17)^. Participants answered on a four-point scale: Strongly agree/Agree/Disagree/Strongly disagree. Responses were categorized as very high, high, low, and very low.

Assessment of COVID-19 vaccine intention and workplace infection control measures To assess vaccine intention, participants were asked: “If a COVID-19 vaccine becomes available, would you like to get it right away?” (yes or no). Workplace infection control measures were evaluated with items covering nine specific measures: prohibition/restriction of business trips; prohibition/restriction of visitors; prohibition of holding or limiting the number of people participating in social gatherings and banquets; restriction on face-to-face meetings; always wear masks during working hours; installation of partitions and change of workplace layout; recommendation for daily temperature check; recommendation for telecommuting; and request not to come to work when sick. These nine items were selected by the researchers based on discussion about infection control measures against COVID-19 in the workplace described in the guidelines of the Japanese government^18)^ and professional organizations^19)^.

### Assessment of covariates

Covariates included demographic and socioeconomic factors, occupation, and number of employees in the workplace. Age was expressed as a continuous variable. Yearly household income was classified into four categories: <2.50 million Japanese yen (JPY); 2.50–3.74 million JPY; 3.75–4.99 million JPY; and ≥5.00 million JPY. Education was classified into four categories: junior high school or high school, vocational school, junior college or technical school, university, and graduate school. Marital status was classified into three categories: married; divorced or bereaved; and unmarried. In the baseline survey, participants chose one of 10 options for their occupation: general employee; manager; executive manager; public employee, faculty member or non-profit organization employee; temporary/contract employee; self-employed; small office/home office; agriculture, forestry, or fishing; professional occupations (e.g., lawyer, tax accountant, medical-related); and other occupations. As noted above, three of these categories were excluded from this study, meaning the occupations were ultimately classified into seven categories. The number of employees in the workplace was classified into four categories: 1–9, 10–99, 100–999, and ≥1000. In addition, the cumulative incidence rate of COVID-19 infection 1 month before the survey was conducted in the prefectures of residence was used as a community-level variable. This information was collected from the websites of public institutions.

### Statistical analyses

The odds ratios (OR) for the association between intention to have the COVID-19 vaccination and POS were estimated using a multilevel logistic model nested in the prefectures of residence to account for area variety. The multivariate model was adjusted for sex and age (Model 1), and then additionally adjusted for equivalent income (categories), educational background (categories), marital status, occupation, and number of employees in the workplace (categories) (Model 2). Finally, the model was additionally adjusted for number of workplace infection control measures (Model 3). All analyses used the incidence rate of COVID-19 by prefecture as a prefecture-level variable. A p-value less than 0.05 was considered statistically significant. All analyses were conducted using Stata (Stata Statistical Software: Release 16; StataCorp LLC, TX, USA).

## Results

Table 1 shows participants’ characteristics by POS category. Of the 23,846 participants, 1,958 (8%) reported very high POS. In very high POS category 836 (43%) workers wanted the COVID-19 vaccination, and in very low POS category 1,382 (36%) workers wanted vaccination.

**Table 1.**
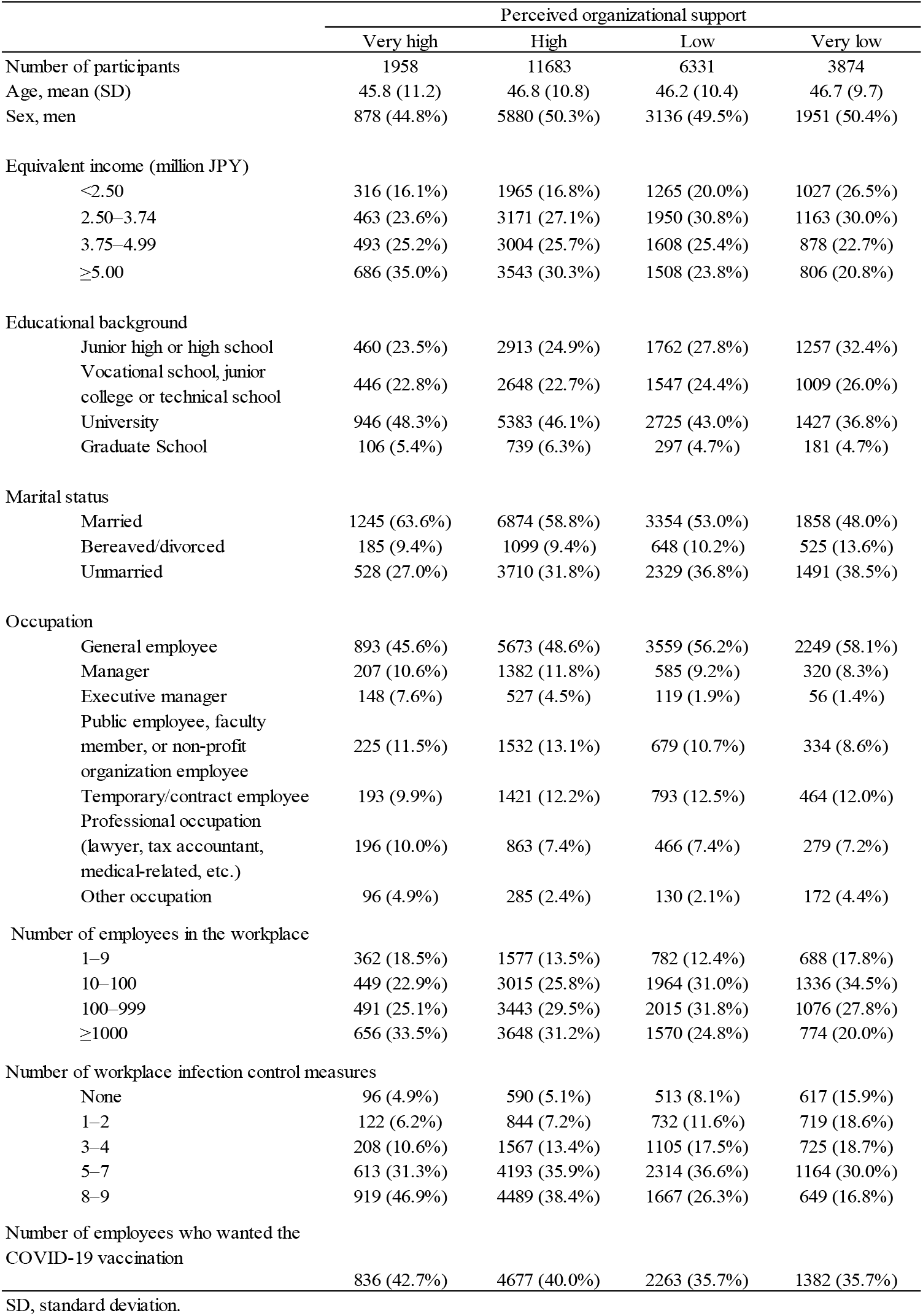
Participants’ characteristics by category of perceived organizational support

Table 2 shows the association between POS (continuous and categories) and COVID-19 vaccine intention. In the analysis with POS as a continuous variable, POS was associated with COVID-19 vaccine intention (Model 2: OR=1.11, 95% confidence interval [CI]: 1.08–1.15, p<0.001). The OR decreased after additional adjustment for number of workplace infection control measures (Model 3: OR=1.06, 95% CI: 1.02–1.09, p<0.001). In the analysis with POS as a categorical variable, very high POS was associated with COVID-19 vaccine intention (reference: very low POS) (Model 2: OR=1.34, 95% CI: 1.20–1.51, p<0.001). The OR decreased after additional adjustment for number of workplace infection control measures (Model 3: OR=1.17, 95% CI: 1.04–1.31, p<0.001). In contrast, high POS was associated with COVID-19 vaccine intention (Model 2: OR=1.17, 95% CI: 1.09–1.27, p<0.001), but was not associated with vaccine intention after additional adjustment for number of workplace infection control measures (Model 3: OR=1.05, 95% CI: 0.97–1.13, p=0.276).

**Table 2.**
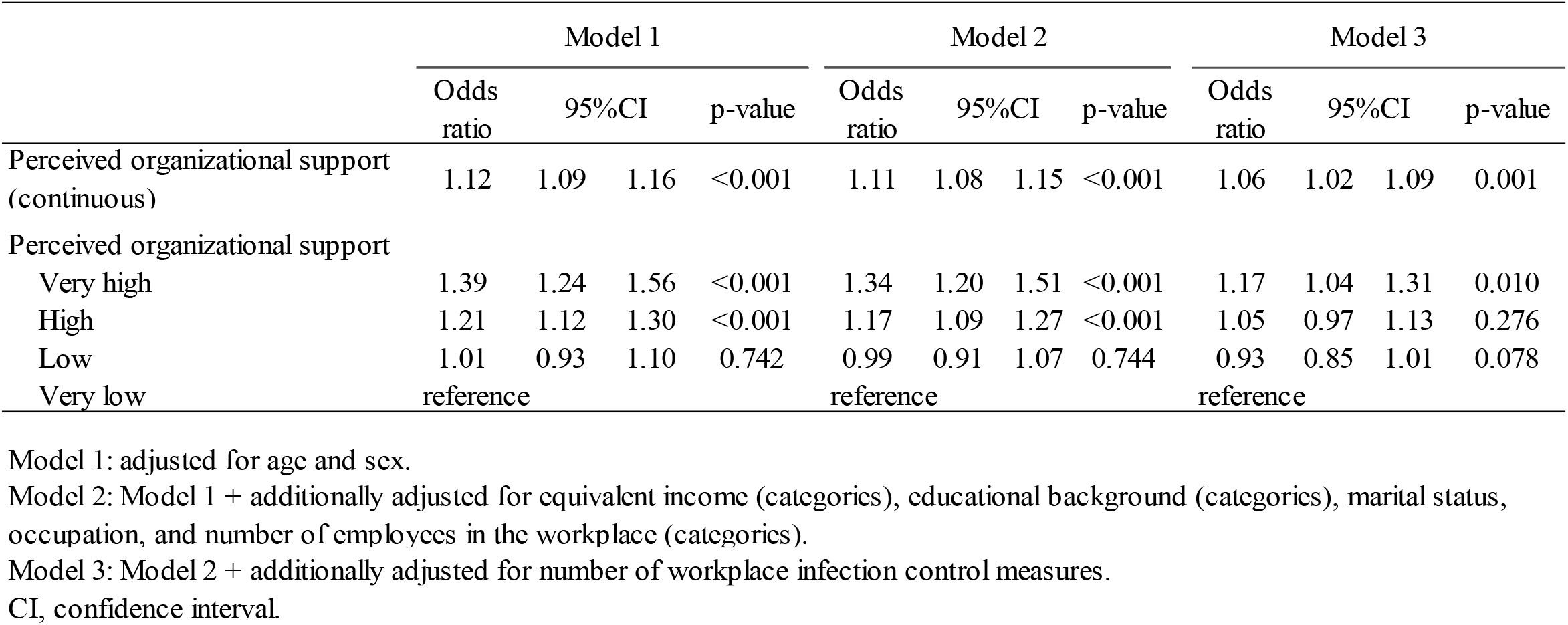
Odds ratios for perceived organizational support (continuous and each category) and COVID-19 vaccine intention in a multiple logistic regression model

## Discussion

This study showed that higher the POS in the population, the higher the vaccine intention. The relationship between POS and vaccine intention was attenuated when adjusted for workplace infection prevention measures. POS is based on the social exchange theory, which suggests that in workplaces with POS, there may be a sense of responsibility toward the group and altruistic behavior toward organizational members, meaning vaccine intentions for infection prevention may have increased^20,21)^. Vaccine intention also includes the social benefits of vaccination. The 5C (confidence, complacency, constraints, calculation, and collective responsibility) scale has previously been used as an indicator to evaluate the complex factors related to vaccine intention. In the 5C scale, collective responsibility is positively correlated with both collectivist tendencies and community orientation and is considered important because it corresponds to the protection of others through herd immunity through one’s own vaccination behavior^22)^.

There are two possible reasons why the association between POS and vaccine intention was attenuated by adjusting for workplace infection prevention measures. First, if workplace infection prevention measures are a confounding factor, proactive infection prevention measures during the COVID-19 pandemic are expected to increase employees’ POS. Although POS does not increase with measures based on laws and regulations or mandatory measures, it is highly valued by organizational members when measures are based on discretionary choices of the organization^23)^. In addition, proactive infection prevention measures may have increased employee health literacy and weakened their concern about vaccines. Conversely, even after adjusting for infection prevention measures, POS was associated with vaccination intention, suggesting that POS itself may have a direct effect on vaccine intention. Second, POS during the COVID-19 pandemic may have facilitated the implementation of workplace infection prevention measures. The presence of leadership support in workplace health promotion has been reported to increase POS^15)^, and high POS in the workplace may increase employee participation in health behaviors^24)^. The same is likely to be true for workplace infection prevention measures that are implemented with a collaborative understanding between employees and the company. As a result, implementation of various workplace infection prevention measures may have increased understanding of vaccines and fostered a “norm of preventing infection” in the workplace, which may have been associated with vaccination intention.

Vaccine intention is based on the relative perception of the efficacy of vaccination and the risk for adverse reactions. As of December 2020, when the survey for this study was conducted, international information regarding vaccine trials and vaccination was still in being received, and it is unlikely that the public had sufficient accurate information on the COVID-19 vaccine. Therefore, it is possible that correct knowledge about the seriousness of infectious diseases gained in the process of implementing workplace infection prevention measures may have raised awareness regarding the need for vaccination. It has also been noted that vaccination behavior is susceptible to social norms, and this may have influenced the intention to vaccinate^25-28)^. POS is rooted in the mutual relationship between the organization and its employees. The present results suggested that a good mutual relationship between the two on a regular basis may lead to an appropriate response to protect the health and life of the organization and its employees in the event of a crisis such as the COVID-19 pandemic. In addition, high POS has been associated with employees’ work-related indicators, such as high organizational commitment, job performance, and lower turnover attitudes^23)^. Conversely, it has been suggested that an organization taking proactive actions to ensure the health and life of its employees during a crisis may have a positive impact on subsequent employee performance.

This study had several limitations that warrant mention. First, the generalizability of the results is uncertain because this study was conducted through an Internet panel. However, we attempted to reduce bias in the target population as much as possible by sampling according to region, job type, and prefecture based on the infection incidence rate. Second, the timing of the survey might have affected the responses of the target population. Since the survey was conducted when Japan was experiencing a full-scale spread of infection, vaccine intention and the status of implementation of workplace infection prevention measures may have been affected. In addition, the vaccination plan in Japan was undecided and it was unclear when the vaccination would be available, which may have influenced vaccine intention responses. Third, POS was evaluated with a simple question (“Your company supports employees in finding a balance between active, productive working and healthy living” (Strongly agree/Agree/Disagree/Strongly disagree), and the measurement validity of the original concept of POS was untested.

This study suggests that high POS during the COVID-19 pandemic increases employees’ vaccination intention. In addition, the relationship between POS and vaccination intention is strongly influenced by implementation of workplace infection prevention measures. Therefore, promoting the improvement of employees’ well-being and implementing appropriate workplace infection prevention measures in the event of an emerging infectious disease outbreak may influence the vaccination behavior of employees. In turn, this may contribute to the acquisition of herd immunity in the community.

## Data Availability

The data that support the findings of this study are available from the corresponding author, TN, upon reasonable request.

## Acknowledgements

This study was supported and partly funded by the University of Occupational and Environmental Health, Japan; General Incorporated Foundation (Anshin Zaidan); The Development of Educational Materials on Mental Health Measures for Managers at Small-sized Enterprises; Health, Labour and Welfare Sciences Research Grants; Comprehensive Research for Women’s Healthcare (H30-josei-ippan-002); Research for the Establishment of an Occupational Health System in Times of Disaster (H30-roudou-ippan-007); and scholarship donations from Chugai Pharmaceutical Co., Ltd., the Collabo-Health Study Group, and Hitachi Systems, Ltd.

Current members of the CORoNaWork Project (in alphabetical order) are: Dr. Yoshihisa Fujino (present chairperson of the study group), Dr. Akira Ogami, Dr. Arisa Harada, Dr. Ayako Hino, Dr. Hajime Ando, Dr. Hisashi Eguchi, Dr. Kazunori Ikegami, Dr. Kei Tokutsu, Dr. Keiji Muramatsu, Dr. Koji Mori, Dr. Kosuke Mafune, Dr. Kyoko Kitagawa, Dr. Masako Nagata, Dr. Mayumi Tsuji, Ms. Ning Liu, Dr. Rie Tanaka, Dr. Ryutaro Matsugaki, Dr. Seiichiro Tateishi, Dr. Shinya Matsuda, Dr. Tomohiro Ishimaru, and Dr. Tomohisa Nagata. All members are affiliated with the University of Occupational and Environmental Health, Japan.

## Ethical approval

This study was approved by the Ethics Committee of the University of Occupational and Environmental Health, Japan (reference No. R2-079 and R3-006).

## Informed Consent

Informed consent was obtained in the form of the website. Registry and the Registration No. of the study/Trial: N/A

## Animal Studies

N/A

## Conflict of Interest

The authors declare no conflicts of interest associated with this manuscript.

## Notes

### Competing Interest Statement

The authors have declared no competing interest.

### Clinical Trial

NA

### Author Declarations

Ethical approval: This study was approved by the Ethics Committee of the University of Occupational and Environmental Health, Japan (reference No. R2-079 and R3-006). Informed Consent: Informed consent was obtained in the form of the website.

